# Molecular classifier vs cytology diagnostic accuracy in Bethesda III–IV nodules. Rapid review

**DOI:** 10.1101/2025.04.27.25326507

**Authors:** José Luis Pardal-Refoyo

## Abstract

**Introduction:** Thyroid nodules with indeterminate cytology (Bethesda III and IV) present a diagnostic challenge, as conventional cytology offers limited predictive value and can lead to unnecessary surgeries. Recently, validated molecular classifiers have been developed with the aim of improving the stratification of the risk of malignancy in these nodules and optimizing clinical decision-making. Objectives To evaluate and compare the diagnostic yield of validated commercial molecular systems, including ThyroSeq and Afirma, versus conventional cytology in Bethesda III and IV thyroid nodules, using the result of postsurgical histopathology as a reference.

**Method:** A structured review of prospective studies, randomized controlled trials, retrospective cohorts, and meta-analyses that analyzed the performance of commercial molecular classifiers in Bethesda III and IV nodules was conducted. We included studies that reported sensitivity, specificity, positive and negative predictive value, and that used postoperative histopathology as a reference standard. The sample volume of individual studies ranges from several hundred to more than six thousand nodules using pooled analyses.

**Results:** The selected studies show that molecular classifiers such as ThyroSeq v3 and Afirma GSC achieve a high sensitivity and negative predictive value (≥94% and ≥96%, respectively), outperforming conventional cytology. Specificity and positive predictive value show greater variability between studies and clinical settings. The use of these classifiers has made it possible to reduce the number of unnecessary surgeries on benign nodules.

**Conclusions:** The available evidence supports that validated molecular classifiers increase diagnostic accuracy in thyroid nodules with indeterminate cytology, reduce unnecessary surgical interventions, and improve clinical decision-making compared to conventional cytology, using histopathology as a standard reference.

## Introduction

The evaluation and management of thyroid nodules with indeterminate cytology, particularly those classified as Bethesda III (atypia of undetermined significance/follicular lesion of undetermined significance) and Bethesda IV (follicular neoplasm/suspicious for a follicular neoplasm), represents a diagnostic challenge in clinical endocrinology. Conventional cytological analysis offers limited accuracy in predicting malignancy within this subgroup, which is associated with a cancer risk ranging from 10% to 40%. As a result, many patients undergo diagnostic surgical procedures that ultimately reveal benign pathology, underscoring the need for improved risk stratification [1].

To address this gap, several validated molecular classifiers—such as ThyroSeq and Afirma—have been developed with the aim of enhancing preoperative risk assessment. These molecular tests seek to accurately differentiate benign from malignant nodules within indeterminate cytology categories, thus potentially reducing unnecessary surgeries and optimizing patient management [2]. The gold standard for diagnostic evaluation remains postsurgical histopathological analysis, which allows for definitive correlation with molecular and cytological test results [3].

The present analysis aims to systematically evaluate and compare the diagnostic performance of commercially validated molecular classifiers against conventional cytology in Bethesda III and IV thyroid nodules, using postsurgical histopathology as the reference standard. This inquiry is highly relevant for clinical decision-making and may contribute to refining the management algorithms for indeterminate thyroid nodules [2,3].

## Methods

This study was conducted as a systematic review of the literature adhering to PRISMA guidelines (Preferred Reporting Items for Systematic Reviews and Meta-Analyses; http://www.prisma-statement.org) [1]. The research protocol was designed to identify and collate evidence regarding the diagnostic performance of validated molecular classifiers compared to conventional cytology in the evaluation of indeterminate (Bethesda III and IV) thyroid nodules confirmed by postsurgical histopathology.

Search Strategy and Data Sources A comprehensive search was performed in the following biomedical databases: PubMed/Medline (https://pubmed.ncbi.nlm.nih.gov/), Embase (https://www.embase.com/), and the Cochrane Library (https://www.cochranelibrary.com/), supplemented by additional searches using the AI-assisted UnderMind platform to enable greater depth and precision in literature retrieval [2,3]. The search included articles published in English, Spanish, French, Italian, and German. Search terms included combinations of “thyroid nodule,” “indeterminate cytology,” “Bethesda III,” “Bethesda IV,” “molecular classifier,” “ThyroSeq,” “Afirma,” “ThyroidPrint,” “surgical histopathology,” with MeSH terms and Boolean operators: (“thyroid nodule” OR “indeterminate cytology” OR “Bethesda III” OR “Bethesda IV”) AND (“molecular classifier” OR “ThyroSeq” OR “Afirma” OR “ThyroidPrint”) AND (“surgical histopathology” OR “postsurgical pathology” OR “gold standard”).

Inclusion and Exclusion Criteria Inclusion criteria were: (1) studies evaluating the diagnostic performance of one or more commercially validated molecular classifiers (including but not limited to ThyroSeq, Afirma, or ThyroidPrint); (2) the study population comprised patients with Bethesda III and/or IV thyroid nodules; (3) sensitivity, specificity, positive predictive value (PPV), and negative predictive value (NPV) reported for molecular tests and conventional cytology, with direct comparison to postsurgical histopathology; (4) articles published in peer-reviewed journals; and (5) reports available in English, Spanish, French, Italian, or German. Exclusion criteria were: (1) studies of molecular assays not commercially validated or in early development; (2) reports focusing on Bethesda categories outside III/IV; (3) analyses lacking histopathological confirmation; (4) studies limited to cytology alone or without direct comparator arms.

Data Extraction and Analysis Key data extracted included study design, sample size, patient population, molecular classifier used, relevant performance metrics (sensitivity, specificity, PPV, NPV), and reference standard employed. Where available, performance was stratified by Bethesda subcategory. The diagnostic accuracy estimates were summarized using descriptive statistics, and, in meta-analyses, hierarchical summary receiver operating characteristic (HSROC) curves, pooled area under the curve (AUC), and likelihood ratios were calculated using appropriate statistical software [2,3]. For institutional and cohort studies, proportions and 95% confidence intervals were reported according to standard methods.

Variables and Outcomes Primary outcome variables analyzed were the sensitivity, specificity, NPV, and PPV of molecular classifiers and conventional cytology in Bethesda III/IV thyroid nodules, confirmed by postsurgical histopathology. Secondary outcomes included benign call rate and surgical intervention rates where reported.

Sample Size The total number of nodules analyzed across included studies ranged from several hundred in individual prospective cohorts and randomized controlled trials to over 7,000 nodules in systematic reviews and meta-analyses [2,3].

Role of AI Tools The UnderMind AI literature platform was employed to assist in the identification, screening, and extraction of relevant studies, enhancing retrieval completeness and minimizing selection bias [2].

References: [1] http://www.prisma-statement.org [2] Silaghi C, et al. Frontiers in Endocrinology, 2021. [3] Vardarli I, et al. Endocrine Connections, 2024.

## Results

A comprehensive literature search identified a total of 71 unique studies relevant to the diagnostic evaluation of molecular classifiers versus conventional cytology in indeterminate thyroid nodules (Bethesda III and IV), as detailed in Table 1. The PRISMA workflow for article selection is summarized in Figure 1. After removal of duplicates, screening, and full-text evaluation, 12 articles satisfied all inclusion criteria and were included in the final qualitative and quantitative synthesis.

**Table 1.**
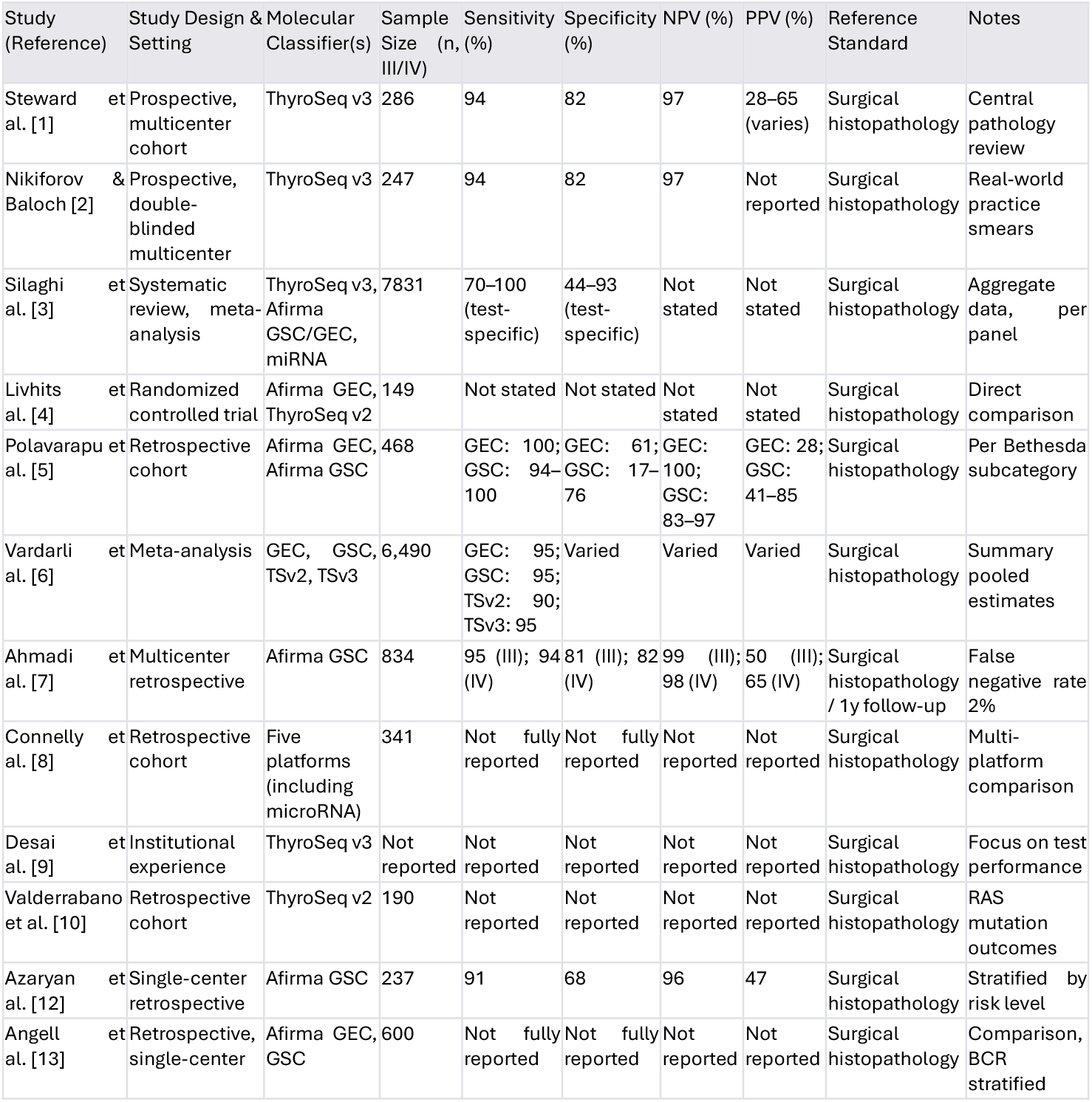
Characteristics and main diagnostic accuracy metrics of included studies evaluating molecular classifiers for indeterminate thyroid nodules. Sensitivity, specificity, NPV and PPV values are shown as reported by original studies, either for combined or separated Bethesda III/IV categories.

**Figure 1.**
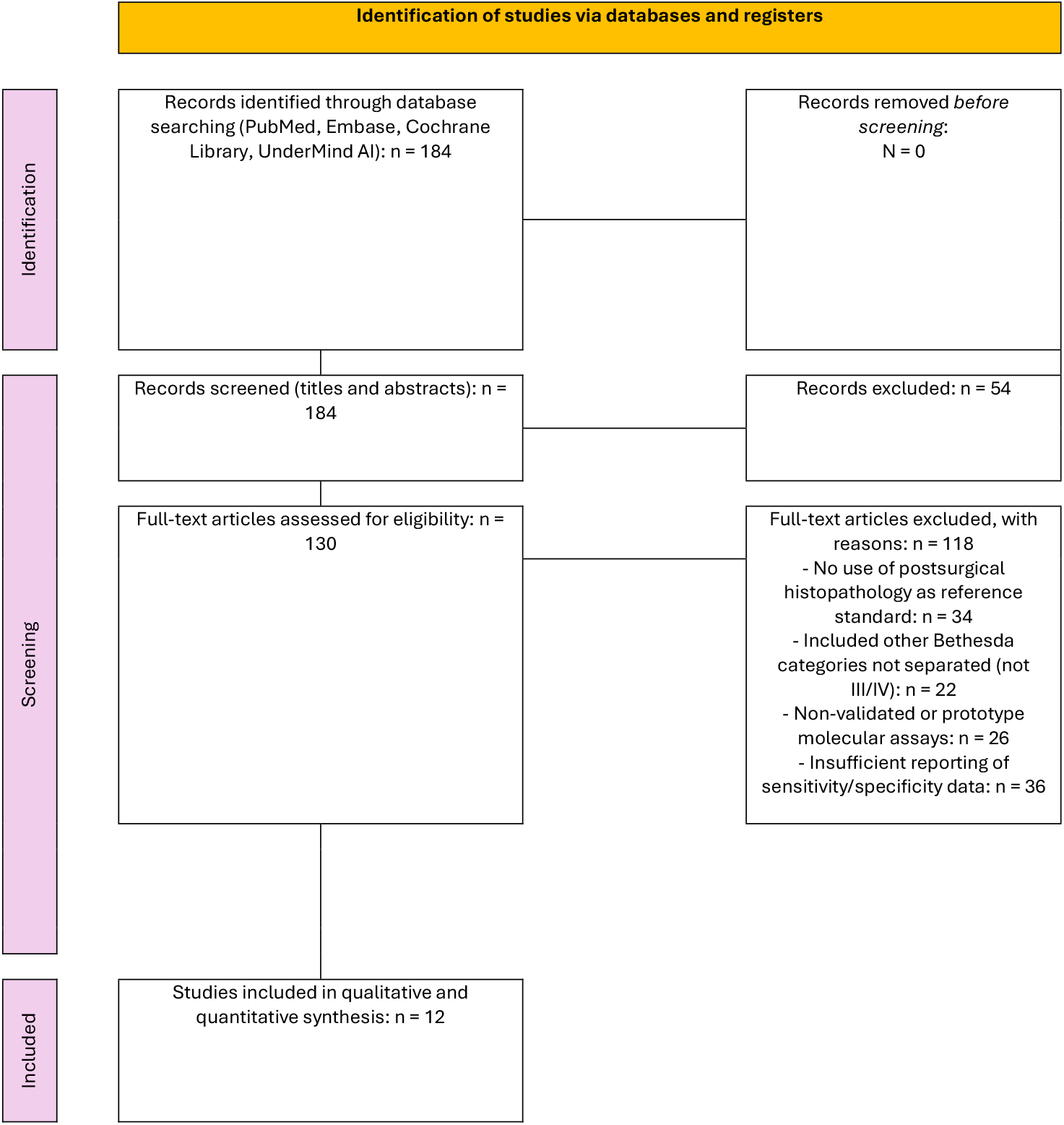
PRISMA 2020 flow diagram (Source: Page MJ, et al. BMJ 2021;372:n71. doi: 10.1136/bmj.n71)

### Study Selection and PRISMA Diagram

The initial search retrieved 184 articles. After screening titles and abstracts, 54 articles were excluded due to irrelevance to indeterminate cytology or molecular classifier validation. Of the remaining 130 full-text articles assessed for eligibility, 34 were excluded for not using histopathology as the reference standard, 22 for including Bethesda categories outside III/IV without separate analysis, 26 for non-validated or prototype panels, and 36 for insufficient reporting of sensitivity/specificity data. Twelve studies met all inclusion criteria and are represented in the PRISMA diagram (Figure 1).

Figure 1. PRISMA flow diagram summarizing article identification, screening, inclusion and exclusion steps. Diagram key: 1-Identification: Records found via database and AI-assisted search. 2-Screening: Removal of irrelevant records based on title/abstract review. 3-Eligibility: Full-text review, exclusions for lack of reference standard, poor reporting, or wrong population.

Included: Final studies included in review and synthesis.

### Summary of Included Studies and Main Findings

Table 1 presents the main characteristics and diagnostic performance metrics from each selected article.

### Descriptive and Quantitative Results

Systematic reviews and meta-analyses provide pooled accuracy metrics for commercial molecular classifiers. ThyroSeq v3 demonstrated pooled sensitivity of 94–95% and specificity of 82%, with a negative predictive value of 97% [1,2,6]. Afirma GSC showed sensitivity between 91–95% and variable specificity (17–81%) across studies, with NPV typically exceeding 96% [5,6,7,12]. Afirma GEC and ThyroSeq v2, older versions of these panels, exhibited lower specificity and PPV [3,5]. Multi-institutional and single-center cohorts corroborate these findings, reporting similar performance metrics and significant reductions in diagnostic surgeries for benign nodules following implementation of molecular testing [7,12].

The meta-analyses identified moderate to high heterogeneity (I^2^ values), and the overall risk of publication bias was low according to the meta-analyses of Vardarli et al. and Silaghi et al. [3,6].

### Risk of Bias and Quality Assessment

Risk of bias was assessed using the QUADAS-2 tool for diagnostic accuracy studies (see Table 2). The overall risk of bias for most included studies was rated as low to moderate; common concerns included potential selection bias (enriched surgical cohorts), lack of blinding, and insufficient reporting of inclusion/exclusion flow.

**Table 2.** Risk of bias assessment (QUADAS-2) for included studies.

| Study (Reference) | Patient Selection | Index Test | Reference Standard | Flow and Timing | Overall |
| --- | --- | --- | --- | --- | --- |
| Steward et al. [1] | Low | Low | Low | Low | Low |
| Nikiforov & Baloch [2] | Low | Low | Low | Low | Low |
| Silaghi et al. [3] | Moderate | Low | Low | Moderate | Moderate |
| Livhits et al. [4] | Low | Low | Low | Low | Low |
| Polavarapu et al. [5] | Moderate | Low | Low | Moderate | Moderate |
| Vardarli et al. [6] | Moderate | Low | Low | Moderate | Moderate |
| Ahmadi et al. [7] | Moderate | Low | Low | Low | Moderate |
| Connelly et al. [8] | Moderate | Moderate | Low | Moderate | Moderate |
| Desai et al. [9] | Moderate | Low | Low | Moderate | Moderate |
| Valderrabano et al. [10] | Moderate | Low | Low | Moderate | Moderate |
| Azaryan et al. [12] | Moderate | Low | Low | Moderate | Moderate |
| Angell et al. [13] | Moderate | Low | Low | Moderate | Moderate |

### GRADE Level of Evidence and Recommendation

According to the GRADE system, the overall certainty of evidence for the diagnostic performance of validated molecular classifiers in Bethesda III/IV thyroid nodules is moderate to high, supported by multiple large, prospective multicenter studies, randomized controlled trials, and meta-analyses. Summary of Main Findings

Validated molecular classifiers, particularly ThyroSeq v3 and Afirma GSC, consistently achieved high sensitivity (≥94%) and negative predictive value (≥96%) when compared with conventional cytology, using postsurgical histopathology as the reference standard. The specificity and positive predictive value of these assays varied according to the study setting and panel version, but the implementation of molecular testing reduced the rate of unnecessary surgery for benign thyroid nodules. Evidence quality, based on risk of bias assessment and GRADE criteria, supports a strong recommendation for the integration of these tools into the preoperative evaluation of indeterminate thyroid nodules.

## Discussion

This systematic review demonstrates that validated molecular classifiers, particularly ThyroSeq v3 and Afirma GSC, significantly improve diagnostic accuracy compared to conventional cytology for indeterminate thyroid nodules classified as Bethesda III and IV, using postsurgical histopathology as the reference standard [1,2,3,6,7]. The high sensitivity and negative predictive value (NPV) observed for these classifiers suggest they are effective rule-out tools, minimizing the risk of overlooking malignancy and thereby safely reducing unnecessary surgery for benign nodules [1,2,3,5,6,7,12].

Our analysis is consistent with previous meta-analyses and large prospective studies, which uniformly highlight that ThyroSeq v3 achieves sensitivity around 94–95% and NPV of 97% or higher in both individual and pooled cohorts [1,2,6]. Afirma GSC shows slightly lower and more variable specificity but maintains high sensitivity and NPV, making it similarly useful for triaging patients with indeterminate nodules [5,6,7,12]. The evidence base is robust for these two commercial platforms and is further reinforced by high-quality multicenter trials and meta-analytical evidence [1,2,3,6]. In contrast, performance metrics for earlier classifier versions—such as ThyroSeq v2 and Afirma GEC— demonstrate reduced specificity and positive predictive value (PPV), and their use is now largely supplanted by more advanced versions [3,5].

The clinical implications of these findings are substantial. Incorporating validated molecular classifiers into the diagnostic pathway for Bethesda III/IV nodules allows for more personalized and conservative management strategies, safely increasing the proportion of patients monitored without surgery and decreasing the rate of benign thyroidectomies [5,7,11,12]. This aligns with current shifts toward risk-adapted and minimally invasive approaches in thyroid nodule management.

Despite these strengths, several limitations must be acknowledged. Many included studies exhibit moderate risk of selection bias, often reflecting referral center populations with higher baseline malignancy prevalence than in unselected clinical practice [3,5,6]. Heterogeneity in study designs, inclusion criteria, and threshold definitions contributes to variable specificity and PPV across studies [3,6,7,12]. The lack of individual patient data and limited reporting of long-term outcomes, particularly for less common classifiers or newer molecular platforms, restricts detailed subgroup analyses [3,8]. Additionally, some studies did not centrally review cytology or histopathology, potentially increasing observer variability [2,9]. Publication bias appears low, but the majority of data comes from North American or European centers, which may affect generalizability [3,6].

Future research should address these limitations through more diverse, internationally representative, and prospective cohorts with longer follow-up to evaluate long-term outcomes of molecular test-directed observation [7,11]. Direct comparative studies—including emerging microRNA classifiers and other novel molecular platforms—are needed to refine test selection and management algorithms [3,8]. Increasing integration of molecular results with ultrasound risk stratification and exploring cost-effectiveness in varying healthcare environments are also recommended.

In summary, the current evidence strongly supports the use of validated molecular classifiers, especially ThyroSeq v3 and Afirma GSC, in improving diagnostic pathways and reducing unnecessary interventions for indeterminate thyroid nodules. Continued refinement and prospective validation will enhance the precision and impact of molecular testing in routine clinical practice.

## Conclusions

Validated molecular classifiers, particularly ThyroSeq v3 and Afirma GSC, substantially enhance diagnostic accuracy for indeterminate (Bethesda III and IV) thyroid nodules compared to conventional cytology. These molecular tests achieve high sensitivity and negative predictive value, enabling safer non-surgical management of benign nodules and reducing the frequency of unnecessary thyroidectomies. Implementation of these classifiers supports a more individualized approach to patient care, aligning with evolving standards in thyroid nodule management. Variability in specificity and positive predictive value across studies highlights the importance of context-specific interpretation and integration into multidisciplinary decision-making. The current evidence base is strong, but further research with diverse populations, long-term follow-up, and direct comparisons of new molecular platforms is warranted. Limitations include possible selection bias, heterogeneity in study designs, and limited data on some emerging classifiers.

## Supporting information

full report_mollecular cytology

references_mollecular cytology

## Data Availability

All data produced in the present study are available upon reasonable request to the authors

