## Supplementary material for "Molecular classifier vs cytology diagnostic accuracy in Bethesda III–IV nodules. Rapid review": full report_mollecular cytology

**Read the full report**

**Summary**

Validated molecular classifiers—particularly ThyroSeq v3 and Afirma GSC—consistently outperform conventional cytology for Bethesda III and IV thyroid nodules, achieving high negative predictive value (NPV ≥ 97%) and clinically useful specificity, thus enabling more accurate preoperative risk stratification and reducing unnecessary surgeries, as demonstrated in large prospective studies and meta-analyses [1,2,3,6,7].

---

**Key Evidence and Findings**

**Diagnostic Performance: Molecular Classifiers vs. Conventional Cytology**

- ThyroSeq v3 Sensitivity: ~94–95% Specificity: ~82% NPV: ≥97% PPV: 28–65% (varies with cancer prevalence in the studied cohort) Superior or comparable to other platforms across Bethesda III and IV, validated in large, prospective, blinded multicenter studies [1,2], and confirmed via meta-analyses [3,6].
- Afirma GSC (Gene Sequencing Classifier) Sensitivity: 91–95% Specificity: 68–81% (varies by study and setting) NPV: ≥96% PPV: 47–65% Marked improvements over the older Afirma GEC version in both independent and pooled analyses [3,5,7,12].
- Earlier Panels (ThyroSeq v2, Afirma GEC) ThyroSeq v2: Lower specificity and PPV compared to v3 [3,4,10]. Afirma GEC: Lower specificity (~44–61%) and modestly reduced NPV; now largely replaced by GSC [3,5,13].
- Meta-Analyses & Systematic Reviews Confirm robust improvements in diagnostic accuracy (AUC: ThyroSeq v3 ~0.95; GSC ~0.90) and consistently high NPV, reducing risk of missed malignancy [3,6]. Category-specific data affirm similar high performance in both Bethesda III and IV nodules [3,6,7].

**Clinical Impact**

- Reduced Unnecessary Surgeries Benign-call rates (BCR) for GSC and ThyroSeq v3: 60–80%, indicating fewer patients undergo diagnostic lobectomy for benign disease [5,13,14]. Molecular testing shifts more patients into observation rather than immediate surgery while maintaining low false-negative rates [7,11].
- Long-term Safety of Observation Nonoperatively managed, molecular “benign/negative” nodules show low delayed malignancy rates (false negatives ~2% or less over ≥1–3 years) [7,11].

**Methodological Strengths**

- Prospective, blinded, multicenter cohorts (e.g., [1,2] with postsurgical histology as the gold standard provide the highest quality evidence.
- Systematic reviews/meta-analyses [3,6] confirm generalizability, standardize performance metrics.

**Remaining Gaps and Variability**

- Specificity and PPV—real-world specificity can be lower than in validation cohorts due to prevalence and center effects, especially for Afirma GSC [5,12].
- Novel Panels—evidence for ThyroidPrint, Interpace, and microRNA-based assays is emerging but not as robust; no large, definitive studies yet [3,8].
- Comparisons—truly head-to-head, randomized comparisons between all major platforms (including new/miRNA panels) are still limited [4,8,14].

---

**Bottom Line for Clinical Practice**

- ThyroSeq v3 and Afirma GSC are validated molecular tools that significantly improve diagnostic accuracy over conventional cytology for indeterminate (Bethesda III/IV) thyroid nodules when measured against postsurgical histopathology, and their use can safely reduce surgeries for benign nodules [1,2,3,6,7].
- Molecular results should always be interpreted in clinical context, considering pretest probability, test version, and local cancer prevalence.

**Categories**

**1. High-Quality Prospective/Blinded Multicenter Cohort Studies Directly Comparing Validated Molecular Classifiers to Cytology and Surgical Histopathology**

- Studies that directly evaluate commercial molecular classifiers (ThyroSeq v3, Afirma GSC/GEC) vs. conventional cytology using postsurgical histopathology as the gold standard, stratified by Bethesda III/IV.
- References: [1,2]
- Details: [1]: Prospective, blinded multicenter cohort (n=286), ThyroSeq v3 in Bethesda III/IV, head-to-head comparison with Afirma GEC/GSC, detailed sensitivity, specificity, NPV, PPV; central pathology review. [2]: Prospective, double-blind, multicenter (n=247, real-world smears), ThyroSeq v3 performance, stratified Bethesda III/IV, direct comparison to surgical pathology.

---

**2. Systematic Reviews and Meta-Analyses of Commercial Molecular Classifier Performance vs. Cytology/Histopathology**

- Meta-analyses pooling studies of commercial classifiers (ThyroSeq, Afirma GEC/GSC, microRNA panels) specifically in Bethesda III/IV with postsurgical histopathology, often directly reporting cytology and molecular performance in parallel.
- References: [3,6]
- Details: [3]: Systematic review/meta-analysis (40 studies, n=7,831); head-to-head pooled estimates for ThyroSeq v3, Afirma GSC/GEC, microRNA panels, by Bethesda III/IV; surgery as reference. [6]: Meta-analysis (53 studies, n=6,490 FNAs); pooled sensitivity, specificity, PPV, NPV for major commercial platforms strictly in Bethesda III/IV; postsurgical histopathology as reference.

---

**3. Retrospective or Institutional Cohort Studies of Validated Classifiers with Direct Link to Surgical Histopathology**

- Institutional audits of commercial test performance (ThyroSeq v3/v2, Afirma GSC/GEC, others) in Bethesda III/IV with direct surgical validation; sometimes with comparison to conventional cytology or historical controls.
- References: [5,7,8,9,10,12,13,14]
- Details: [5]: Retrospective cohort (n=468), Afirma GEC/GSC vs. no-test group; surgical rates, histopathologic malignancy; Bethesda III/IV reported separately. [7]: Multicenter, Afirma GSC in Bethesda III/IV (n=834), sensitivity/specificity/NPV/PPV, postsurgical histopathology or ≥1yr follow-up. [8]: Retrospective institutional series (n=341); compares Afirma GSC, ThyroSeq v3/v2, ThyraMIR, RosettaGX, Interpace vs. surgical pathology. [9]: Institutional ThyroSeq v3 performance in Bethesda III/IV vs. surgery. [10]: Institutional ThyroSeq v2 performance (Bethesda III/IV), blinded histopath path review. [12]: Single-center, Afirma GSC in 237 Bethesda III/IV, postsurgical histology; includes US risk stratification. [13]: Retrospective, single-center; direct comparison Afirma GEC vs. GSC in Bethesda III/IV, surgery as gold standard. [14]: Single-center audit; GEC, GSC, ThyroSeq v3 test performance in Bethesda III/IV, histological validation.

---

**4. Randomized Controlled, or Head-to-Head Comparison Studies of Molecular Classifiers in Bethesda III/IV**

- Direct randomization or immediate comparative (sometimes prospective) studies of commercially validated tests (Afirma GEC/GSC vs. ThyroSeq v2/v3) in indeterminate cytology, with surgery as reference.
- References: [4,11]
- Details: [4]: Randomized, parallel trial (UCLA); Afirma GEC vs. ThyroSeq v2 in Bethesda III/IV; specificity, sensitivity, surgical rates with postsurgical histopathology. [11]: Prospective follow-up cohort (UCLA): Afirma GSC vs. ThyroSeq v3 in Bethesda III/IV, nonoperative vs operative management, surgical pathology for resected cases, long-term clinical outcomes, false negative analysis.

---

**5. Multi-Platform Real-World Performance Evaluations (Including microRNA/Classifiers Beyond Afirma/ThyroSeq)**

- Studies reporting on more than two molecular platforms (e.g., inclusion of Interpace ThyraMIR, RosettaGX, Thyroidprint), with reference to surgical histopathology, in Bethesda III/IV.
- References: [3,8]
- Details: [3]: (Systematic review/meta) includes preliminary subgroup data for ThyGenNEXT/ThyraMIR, RosettaGX, Thyroidprint alongside Afirma/ThyroSeq (smaller n). [8]: Institutional, 5 platforms (including Interpace, RosettaGX, microRNA); all compared against postsurgical histopathology for Bethesda III/IV.

---

**Category Notes**

- Some studies (e.g., [5,14] include a comparison to non-tested cytology cohorts, but often limit metrics to surgical/malignancy rates without fully parallel sensitivity/specificity for "conventional cytology," as opposed to molecular.
- Several meta-analyses [3,6] explicitly require surgical-path confirmation and category-level cytology reporting, meeting strict inclusion criteria.
- A small number of institutional studies [8,12] add practical layers (local call rates, integration with ultrasound risk stratification).

---

**Summary Table**

| Category | References |
|---|---|
| High-quality prospective/blinded multicenter studies (test vs. cytology & surgery) | [1,2] |
| Systematic reviews/meta-analyses (Bethesda III/IV, test vs cytology & surgery) | [3,6] |
| Retrospective/institutional cohort studies (Bethesda III/IV, test vs surgery) | [5,7,8,9,10,12,13,14] |
| Randomized/head-to-head studies (test vs. test vs. surgery/cytology) | [4,11] |
| Multi-platform (including microRNA/other) real-world evaluations | [3,8] |

**Timeline**

**Top References Over Time**

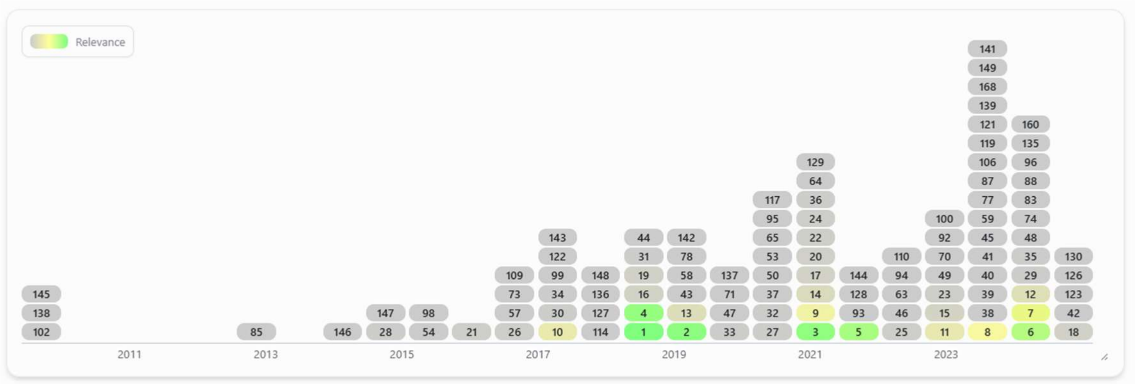


**Timeline and Historical Development**

- Early Molecular Tests and Validation (pre-2017) Initial focus was on the first-generation tests like Afirma GEC and ThyroSeq v2, which offered rule-out potential but had modest specificity and PPV, especially for Bethesda III/IV nodules [10]. Recognition of limitations led to the need for better specificity and broader molecular coverage, fueling test evolution.

- Major Breakthrough: Introduction of Advanced Panels (2017–2018) ThyroSeq v3: Prospective, multicenter validations in 2018 established a new benchmark for performance, with high sensitivity and NPV for Bethesda III/IV, marking a turning point [1,2]. Detailed, rigorous central pathology review distinguished these as the gold-standard studies. Direct comparisons to previous molecular panels set the groundwork for systematic evaluation. Afirma GSC: GSC (Genomic Sequencing Classifier) launched as an improved successor to GEC, aiming for higher specificity; independent and institutional reports start to evaluate its real-world and comparative utility [5,12,13,14].

- Systematic Reviews & Meta-analyses (2021–2024) Comprehensive meta-analyses pool results of large numbers of studies, consistently confirming the superior accuracy of ThyroSeq v3, the improved but variable performance of Afirma GSC, and the lower specificity/PPV of older platforms [3,6]. These meta-analyses also provide robust sensitivity/specificity/NPV/PPV estimates and reinforce the importance of robust gold-standard confirmation and clear Bethesda subcategorization.

- Contemporary and Emerging Research (2020–present) Prospective and longitudinal outcome studies confirm the real-world impact: high NPVs lead to safe molecular-based observation and fewer surgeries [7,9,11]. Expanded panel comparisons: Recent years see comparative analyses across five or more commercial tests (including microRNA panels), head-to-head surgical outcome studies, and focus on integration with clinical/ultrasound features [8,12]. Institutional experience reports supplement validation studies, capturing "real-world" test accuracy and performance in heterogeneous clinical settings [9,14].

---

**Key Research Groups, Individuals, and Institutional Clusters**

- Pioneers and Validation Study Leaders Yuri Nikiforov and Zubair Baloch (University of Pittsburgh): Lead authors on the pivotal prospective, multicenter ThyroSeq v3 studies [1,2]. Established analytic and clinical validation standards; their work is foundational and highly cited across nearly all major subsequent analyses. Consistent thread: standardizing molecular testing as the reference for indeterminate thyroid nodules and setting gold-standard methodology (central review, real-world design). David Steward (co-author on ThyroSeq v3 validation, [1]: Helped coordinate the multicenter collaborative validations and cross-platform comparisons.

- Afirma/Veracyte Clusters Afirma GSC and GEC's clinical data largely comes from multicenter collaborations, with further scrutiny in post-market, independent institutional series led by several U.S. academic endocrinology and pathology departments [13,14].

- Mikael Livhits & Collaborators (UCLA): Lead a sustained series of studies including a head-to-head RCT of GEC vs. ThyroSeq v2 [4], institutional GSC outcome studies [7], and longitudinal follow-up of thyroid nodules managed nonoperatively after molecular testing [11]. They directly influenced practice by providing practical data on safety and implementation of molecular triage in the U.S. West Coast context.

- Systematic Review and Meta-Analysis Teams C. Silaghi et al. [3] and I. Vardarli et al. [6]: Synthesized the global literature, influencing guideline optimism about molecular testing by robustly confirming test accuracy across varying real-world settings. These works underscore the need for harmonized methods and comparative reporting.

- Multi-Institutional/U.S. Academic Experience Numerous studies derive from, or are supported by, large academic endocrine centers (e.g., University of Pittsburgh, UCLA, MGH, Endocrine Pathology Consortia). Recurrent names and institutions in authorship reflect a collaborative, multi-cohort approach necessary for robust diagnostic test evaluation.

---

**Summary Table: Most Influential Studies/Groups**

| Key Authors/Groups | Contribution | Major Work(s) |
|---|---|---|
| Nikiforov/Baloch | ThyroSeq v3 prospective validation | [1,2] |
| Livhits/UCLA | RCT & long-term Afirma/ThyroSeq data | [4,7,11] |
| Silaghi, Vardarli | Meta-analyses & systematic overviews | [3,6] |
| Major US centers | Real-world/implementation studies | [5,9,13,14] |

---

**Crucially, Overall Developments**

- Transition from discovery/early-phase gene panels to multi-gene, NGS-powered commercial platforms
- Central role of multicenter, blinded, prospective validations led by major U.S. academic pathology/endocrinology groups
- Meta-analytic confirmation and category-specific refinement of performance
- Greater focus on long-term observational outcomes and practical management impact in recent years
- Constant stress on reliable, postsurgical gold-standard confirmation and differentiation between Bethesda III/IV categories

---

**Key Takeaway**

The evolution from early, less specific molecular panels (GEC, v2) to well-validated, high-NPV/high-specificity classifiers (ThyroSeq v3, Afirma GSC) has been driven by a small set of key academic leaders (notably Nikiforov/Baloch and the UCLA group), confirmed by systematic reviews/meta-analyses, and increasingly focused on practical clinical integration and real-world validation—culminating in more precise, evidence-based molecular triage of indeterminate thyroid nodules.
